# Analysis of Epidemiology and Drug Resistance Patterns of ESKAPE and Non-ESKAPE pathogens at Nigde Hospital in Turkey: A Retrospective Study (2022–2024)

**DOI:** 10.1101/2024.12.19.24318901

**Authors:** Mohammed A. Salim, Songül Budak Diler, Ramazan Köklü, Fikriye Polat, Nooh Mohamed Hajhamed, Ali Özturk

## Abstract

**Purpose:** This study, aimed at determining the epidemiology and antimicrobial susceptibility patterns of infectious diseases caused by ESKAPE and non-ESKAPE microorganisms in patients hospitalized at the Nigde Ömer Halisdemir University Training and Research Hospital in Nigde, Turkey, is a crucial step in understanding and combating the global public health problem posed by the ESKAPE pathogens.

**Materials and methods:** A retrospective analysis was conducted at a primary tertiary care teaching reference hospital in Nigde. The study included patients admitted to the hospital between June 2022 and June 2024. The hospital information system offered clinical and demographic data for the patients. Furthermore, the hospital’s microbiology lab acquired information on bacterial isolates and antibiotic resistance.

**Results:** This study included 13378 bacterial isolates, predominantly gram-negative bacteria, particularly those from the Enterobacterales group. Among these isolates, 9798 (73.2%) were identified as ESKAPE pathogens, and 3580 (26.8%) were identified as non-ESKAPE bacteria. The Intensive Care Unit (ICU) accounted for the highest proportion of infections (34.47%), followed by the pediatric unit (22.6%). The most common infections were caused by *Escherichia coli* (4747 isolates, 35.5%), *K. pneumoniae* (1921 isolates, 19.6 %), and *Acinetobacter baumannii* (1049 isolates, 10.7 %). Furthermore, the analysis revealed that approximately 50.86% of the ESKAPE isolates were classified as multidrug-resistant (MDR) or extensively drug-resistant (XDR). XDR was predominantly detected in *Acinetobacter baumannii* (72.4%), whereas MDR was predominantly detected in *Enterococcus faecium* (76.9%). In contrast, Non MDR was predominantly detected across non-ESKAPE pathogens in *Staphylococcus hemolyticus* (96.1%), *Staphylococcus epidermidis* (86.5%), and *Staphylococcus hominis* (84.5%). Demographic data from the study highlighted significant age group disparities in individuals infected by the ESKAPE and the non-ESKAPE bacteria, with a more substantial proportion of older and children individuals represented in the research sample.

**Conclusions:** This study underscores the significant threat posed by multidrug-resistant ESKAPE pathogens in reference hospital settings, emphasizing the urgent need for effective surveillance and control measures.

## Introduction

Antimicrobial resistance (AMR) poses a significant threat to global public health, jeopardizing our ability to combat infectious diseases effectively [1]. The lack of comprehensive information regarding the prevalence and associated consequences of multidrug-resistant (MDR) diseases further highlights the severity of this issue. Nevertheless, numerous studies have established a clear connection between antibiotic-resistant bacterial infections and patient adverse outcomes. These outcomes include prolonged hospital stays, heightened rates of morbidity, and elevated mortality rates [2–4]. Antibiotic resistance is increasing worldwide, making it difficult to treat bacterial infections, especially those caused by gram-negative bacteria. Limited treatment options exist for multi drugs resistant *Enterococcus faecium* (*E. faecium), Staphylococcus aureus (S. aureus), Klebsiella pneumoniae (K. pneumoniae), Acinetobacter baumannii (A. baumannii), Pseudomonas Aeruginosa (P. aeruginosa), and Enterobacter spp*. (ESKAPE) pathogens cause most nosocomial infections and require urgent antibiotics [5, 6]. The ESKAPE pathogens contribute significantly to nosocomial infections, particularly in critically ill and immunocompromised patients. These multidrug-resistant organisms can avoid the killing effects of antibiotics [7]. Treatment for infections caused by MDR bacteria frequently requires costlier or unsafe therapies (e.g., colistin, daptomycin) [8]. Specific antibiotic resistance in the Enterobacterales family is listed as one of the critical priorities in the World Health Organization’s (WHO) recently developed global priority list of antibiotic-resistant pathogens, which is intended to guide the discovery of new antibiotics (e.g., third-generation cephalosporin-resistant Enterobacterales) [9]. The rise of AMR presents a growing challenge, impacting not only hospitals but also infections acquired in the community. This is linked to the effective infiltration and dissemination of MDR clones from healthcare settings [10]. The global use of antimicrobial drugs in human medicine has increased, particularly in low- and middle-income nations. Nonetheless, these areas still need to catch up to wealthier nations regarding overall usage [11]. Nevertheless, the widespread use of antimicrobials in animals casts a shadow, as it accounts for 80% of antimicrobial consumption in the United States and surpasses 63,000 tons globally, primarily utilized for nontherapeutic reasons such as promoting growth [12].

The prevalence and outcomes of MDR infections associated with the effective spread of MDR clones from hospital-acquired settings into community-acquired infections must be fully comprehended.

## Materials and methods

### Study design and setting

This retrospective study was conducted at Nigde Ömer Halisdemir University Training and Research Hospital, the reference tertiary-care teaching hospital in Nigde, Turkey, from June 2022 to June 2024. The hospital boasts approximately 1000 beds; moreover, the central laboratory microbiology laboratory at the hospital is responsible for bacterial isolation and antimicrobial susceptibility profiles for patients from this hospital and other hospitals or medical health centers in the city. The Republic of Turkey Ministry of Health (RTMH) supervised and sponsored the laboratory. The RTMH provides mentoring and monthly technical support, including external quality control.

Physicians or nurses collected samples from all inpatient and outpatient departments and immediately forwarded them to the microbiological laboratory. The clinicians decided to collect samples for microbial culture and selected the samples themselves. Patient-specific data (age and sex) and a complete record of bacteriological culture and antibiotic resistance profile were gathered from Microbiology Laboratory Information System (LIS) records.

### Study participants and data collection

The study included participants who visited Nigde Ömer Halisdemir University Training and Research Hospital and reported bacterial infections during the study period. This study used a standard data collection form to retrieve all documented data from the microbiology laboratory registration books at Nigde tertiary-care teaching hospital, including patient age and gender, bacterial isolates, and antimicrobial susceptibility profiles.

All laboratory procedures followed Clinical and Laboratory Standards Institute (CLSI) norms and regulations and our microbiology unit’s local standard operating procedures (SOPs). Data acquired during the investigation were kept anonymous and used only for this study. We included all documented data from the study period but excluded illegible or incompletely documented cultures.

### Bacteriological investigation

Our bacteriological analysis follows CLSI recommendations. Clinical samples have been collected from different hospital departments, with the Intensive Care Unit (ICU) and pediatric department being the most infected. The samples were subsequently sent to the laboratory, where established protocols were used to process them. Clinical samples were cultured, including urine, blood, tracheal aspirate, sputum, and swabs from various body sites (rectal and wound). Each clinical sample used standard microbiological culture techniques. Briefly, specimens were collected, inoculated onto suitable isolation culture media (blood agar, chocolate, and eosin-methylene blue (EMB) agar), and incubated at 35-37 °C following established techniques for each sample. Bacterial identification was based primarily on colony features and the Gram-stain reaction, followed by the VITEK®2 compact system (bio-Mérieux) using GN and GP ID cards by CLSI guidelines and the created SOP [13]. The antimicrobial susceptibility profiles of gram-negative and gram-positive bacteria were performed per the CLSI guidelines using the automated VITEK® 2 Compact system (bio-Mérieux). On the other hand, Antimicrobial susceptibility testing (AST) was also performed via the Kirby–Bauer disc diffusion method to determine the antibiotic susceptibility of the bacterial isolates. Antimicrobial susceptibility testing was conducted via the disk diffusion method according to the EUCAST guidelines, and zone diameters were interpreted using the EUCAST clinical breakpoints version 4 (http://www.eucast.org/clinical breakpoints/)[14]. Furthermore, the VITEK®2Compact system (bio-Mérieux) was utilized to establish the minimum inhibitory concentrations (MICs) for antimicrobial susceptibility testing. The results were interpreted using the CLSI criterion [7]. This study defines Multidrug-Resistant Bacteria (MDR) as bacteria resistant to at least one antimicrobial therapy from three different groups. While extensively drug-resistant (XDR) bacteria are only susceptible to one or two antimicrobials, pan-drug-resistant (PDR) microorganisms are resistant to all antimicrobial therapies.

### Statistical analyses Descriptive statistical analysis

Microsoft Excel 2020 was used to complete the analysis (containing means or medians with ranges and percentages to characterize the data). R (R Foundation for Statistical Computing, version 4.4.1) software was used to analyze and visualize the data. The Chi-square or Fisher’s exact test further determined temporal changes in age distributions and AMR. Statistical significance was confirmed if a two-tailed P value was no more than 0.05. Results Alterations of ESKAPE distribution.

### Ethics Approval and consent to participate

The Ethics Committee of the “Ethics Committee for Non-Interventional Clinical Research Nigde Omer Halisdemir University” approved this study with decision number (2022/82), dated 10.09.2022. Given that this was a retrospective study, the Ethics Committee for Non-Interventional Clinical Research Nigde Omer Halisdemir University waived the need for informed consent.

### Results Demographic data

The dataset’s demographic distribution consists of a range of age values, with a minimum age of 0.00, a median of 59.62, an average of 48.13, a majority below 76.02, and a maximum of 101.8, with outliers and higher values. The study revealed 57.6% females and 42.4% males, with a slightly more significant percentage of men; the population was divided into six age groups with predominantly older adults group: (pediatric from 0-5, school-age children from 6-17, young adults from 18-40, middle-aged adults from 41-60, older adults from 61-80 and elderly aged over than 80 years). Additionally, there were significant differences (P < 0.05) in the age groups of patients infected by the ESKAPE and other non-ESKAPE bacteria (Fig 1).

**Fig 1.**
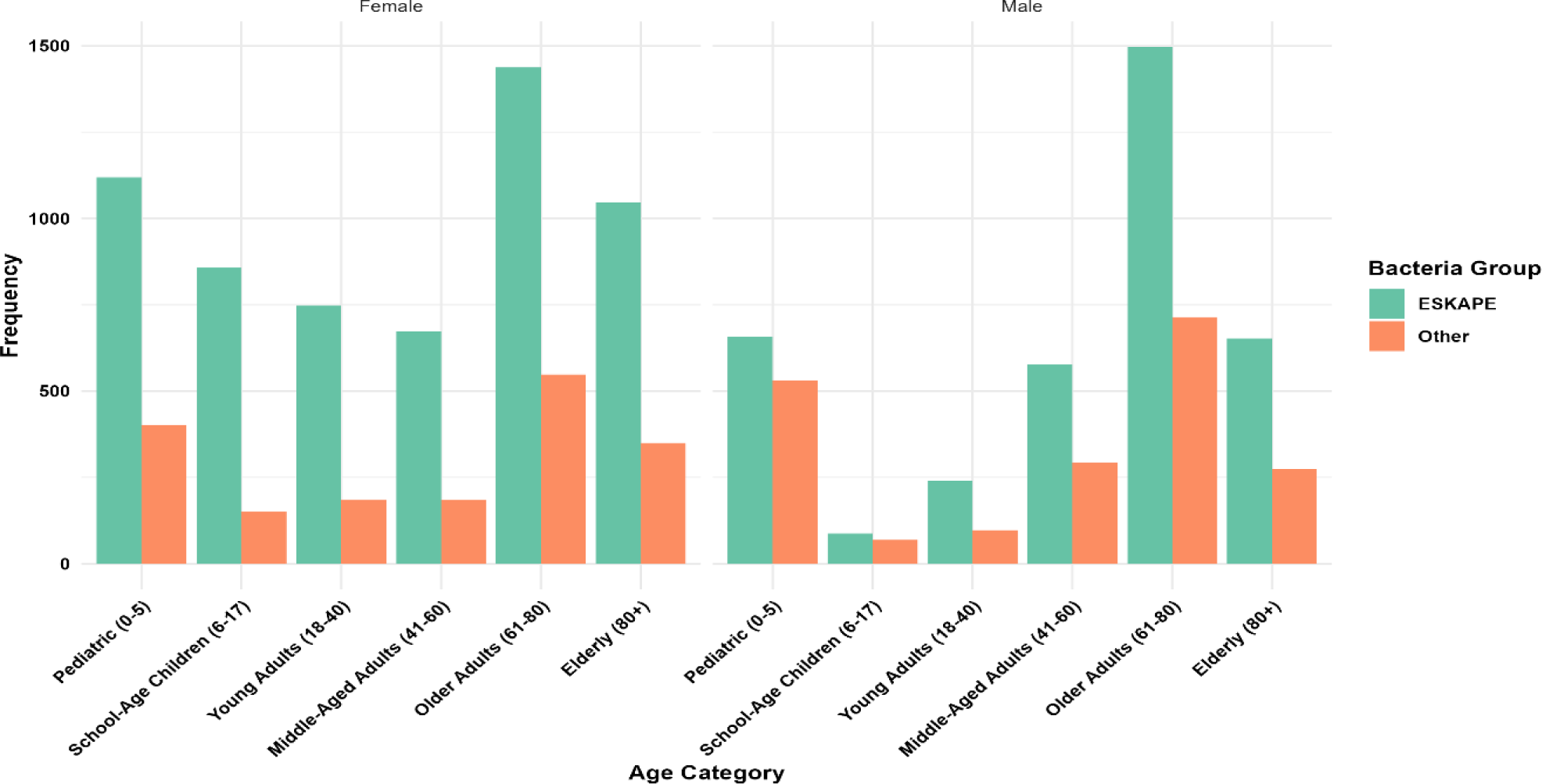
Age groups and frequency of ESKAPE Non_ESKAPE bacterial infection in gender patients.

### Distribution of Bacterial Isolates

During the study period, n = 13378 bacterial isolates were recovered from clinical specimens, of which n = 9798 (73.2%) were ESKAPE pathogens. As shown in Table 1, the study revealed the dominance of gram-negative bacteria. The Enterobacterales group was the predominant antibiotic-resistant bacteria among all the ESKAPE isolates shown in Table 1, with 75% of all the isolates being from this group. *E. coli* was the most frequently isolated bacteria (4747, 35.5%), followed by *K. pneumoniae* (1921; 14.3%). The third most common isolate was *A. baumannii* (1049, 7.8%), followed by *P. aeruginosa* (751, 5.6%). *E. faecalis, S. aureus*, *Staphylococcus hominis (S. hominis), Staphylococcus epidermidis (S. epidermidis),* and *Proteus mirabilis (P. mirabilis)* played limited roles, withless than a 5% share within the ESKAPE group (**Table 1**).

**Table 1.** Distribution and frequency of pathogens during the study period.

| Gram Stain | Bacteria | General<br>Frequency (%) in<br>All isolate<br>13378 | ESKAPE Frequency (%)<br>9798 (73.2%) |  |
| --- | --- | --- | --- | --- |
| gram-negative | *K. pneumoniae* | 1921(14.3%) | 19.6% | 8494<br>(86.6%) |
| ESKAPE | *A. baumannii* | 1049 (7.8%) | 10.7% |  |
|  | *P. aeruginosa* | 751 (5.6%) | 7.6% |  |
|  | *E. coli* | 4747(35.5%) | 48.4% |  |
|  | *E. cloacae* | 207 (1.5%) | 2.1% |  |
| Non-ESKAPE | *P. mirabilis* | 416 (3.1%) | 570 (15% ) |  |
|  | *K.Oxytoca* | 154 (1.2%) |  |  |
| Total |  | 9245 (69.1%) |  |  |
| gram-positive | Total | 3267 | gram-positive ESKAPE<br>1123 (11.5%) |  |
| ESKAPE | *E. faecium* | 542 (4.01%) | 48.2% | 1223<br>(34.4 %) |
|  | *S. aureus* | 581 (4.3%) | 51.73% |  |
| Non-ESKAPE | *E. faecalis* | 606 (4.5%) | 2144(59.7%) |  |
|  | *S. epidermidis* | 571 (4.2%) |  |  |
|  | *S.haemolyticus* | 254 (1.9%) |  |  |
|  | *S. hominis* | 566 (4.2%) |  |  |
|  | *S. agalactiae* | 78 (0.6%) |  |  |
|  | *S. pneumoniae* | 69 (0.5%) |  |  |
| Other |  | 875 (6.5%) | 875(24.3%) |  |

The most common clinical samples included midstream urine or urinary catheter samples, blood cultures, tracheal aspirates, sputum, and wound or abscess samples (**Fig 2**). Rectal swabs, synovial fluid, and stool samples were seldom collected, primarily from seriously ill individuals. *E. coli* were dominated pathogen isolated from urine sample followed by *K. pneumoniae*. Whereas *S. aureus* was most commonly isolated from wound/abscess samples, followed by *A. baumannii* Interestingly, among non ESKAPE bacteria *Staphylococcus hemolytic* were dominated isolates from the blood culture samples where *K. pneumoniae* was the most common ESKAPE strain isolates from the blood culture (**Fig 2**). And the distribution of non-ESKAPE bacteria isolated from different clinical samples (**Supplementary Fig1**).

**Fig 2.**
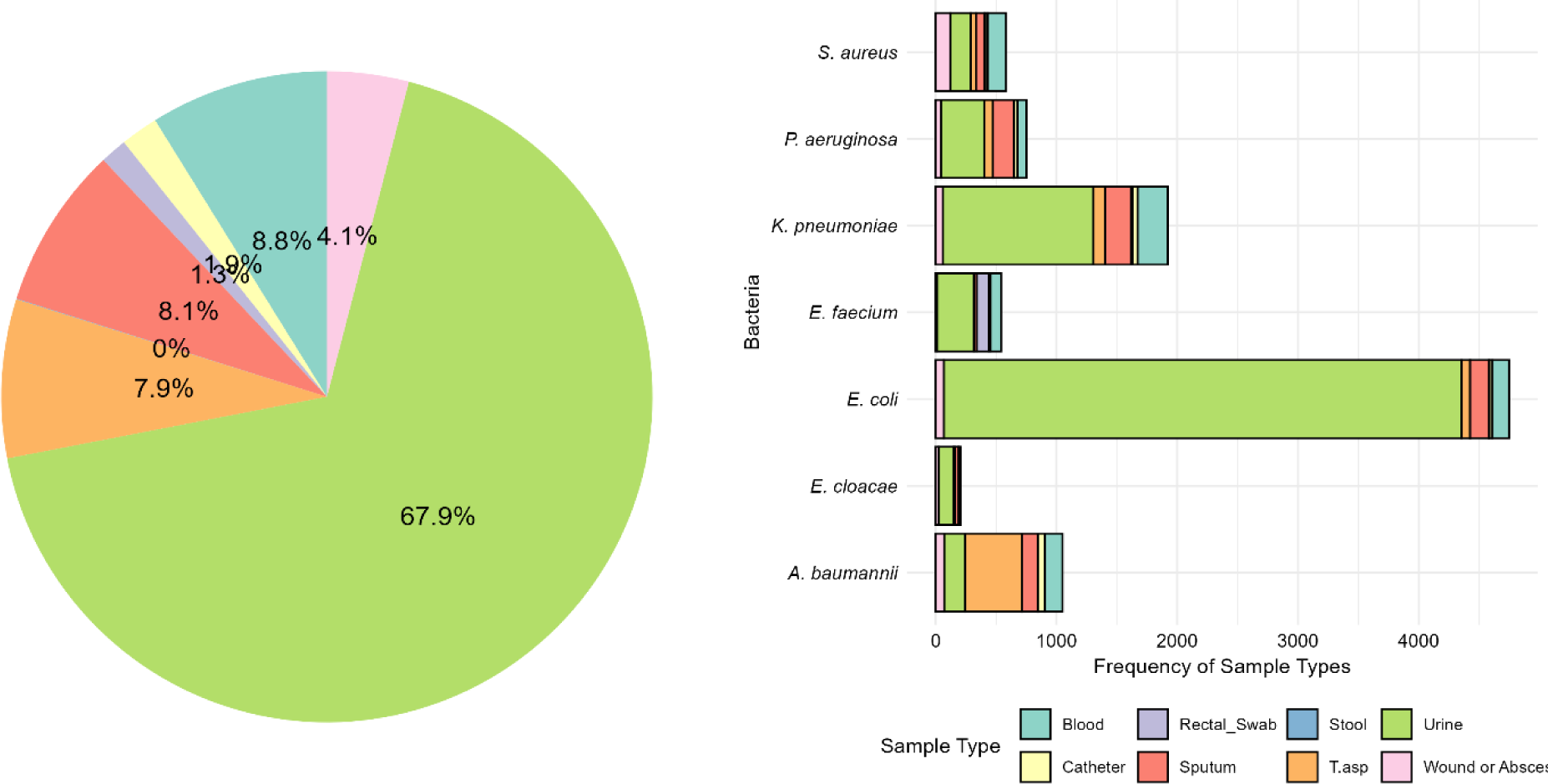
distribution of ESKAPE bacteria isolated across different clinical samples.

In the context of clinical departments, the intensive care unit (ICU) stands out as a significant focal point for healthcare-associated infections. It has been identified as the main source for the colonization and infection of both ESKAPE and non-ESKAPE pathogens, with a notable prevalence of *E. coli, A. baumannii,* and *K. pneumoniae*, being isolated within the ICU environment at a rate of 24%. The ESKAPE bacteria were prevalent in the ICU at 67.7%, requiring special monitoring to ensure patient safety. In pediatric clinics, *E. coli* was found at a prevalence of 40%, accounting for about 20% of all isolated bacteria followed by *K. pneumoniae* (Fig2). This high prevalence necessitates infection control measures for *E. coli* in pediatric clinics. as well as in the urology department *E. coli* was the most commonly isolated in the urology department Detailed data for other bacteria can be found in Supplementary Table 3.

**Fig 2.**
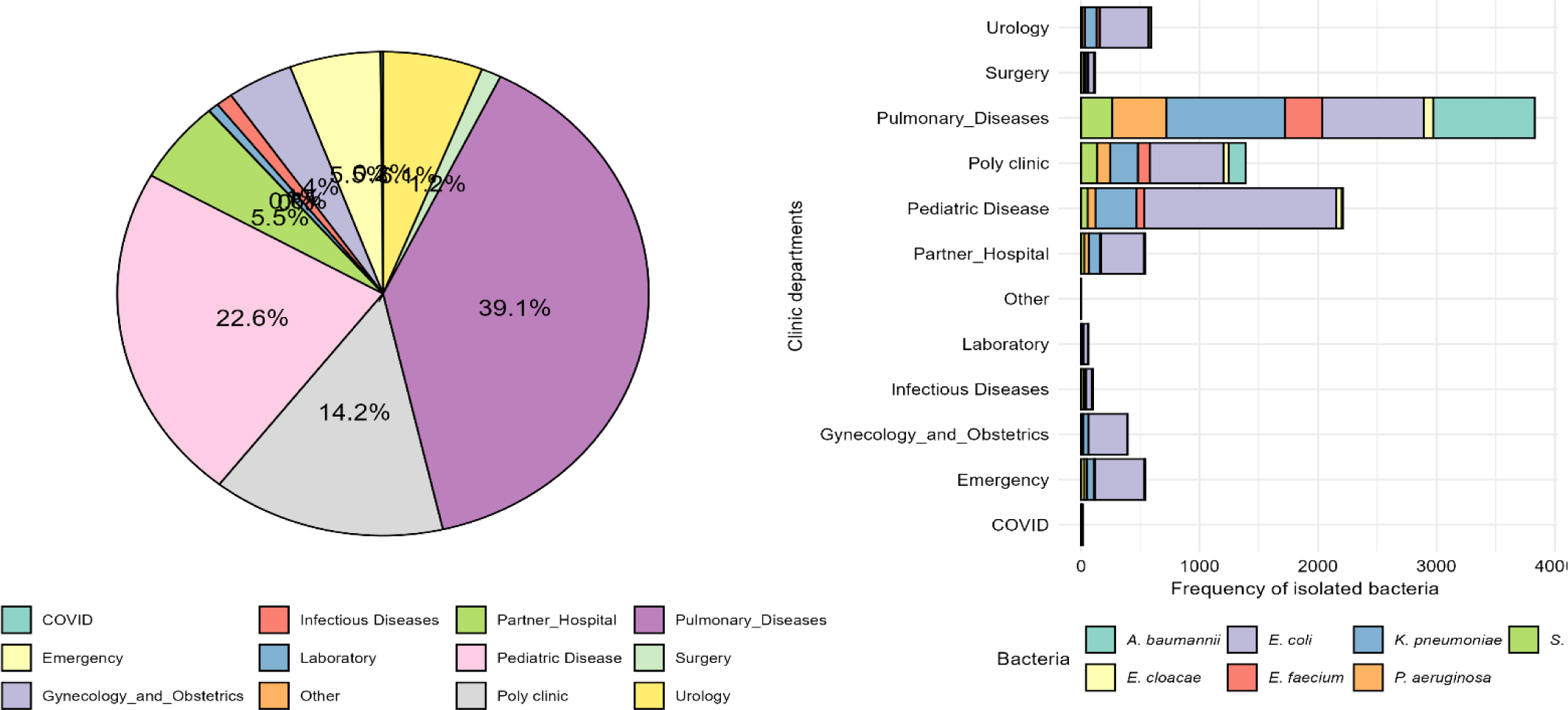
Prevalence of the ESKAPE bacteria across clinical departments.

### Categorization of the ESKAPE pathogens according to the antibiotic susceptibility test

The total resistance level of the ESKAPE isolates has been summarized (**Fig 3**). In the context of resistant phenotype, 49.9% of the isolates were determined to exhibit resistance to at least one antimicrobial therapy from three different groups, with (39.6%) classified as MDR and (10.3%) as XDR. MDR was found in a broad spectrum of ESKAPE isolates, with *E. coli* being the most common, followed by K. *pneumoniae*, *E. faecium A. baumannii*, *S. aureus* and *P. Aeruginosa* and *E. cloacae*. While A. *baumannii* had the highest prevalence of XDR, followed by *K. pneumoniae*, *P aeruginosa*, *E. coli* and *E. faecium,* and no XDR was reported in *S. aureus* and *E. cloacae*. (**Fig 3**). in contrast, the antibiotic susceptibility pattern of the non-ESKAPE bacterial isolate was seen in **Supplementary Fig 2**.

**Fig 3.**
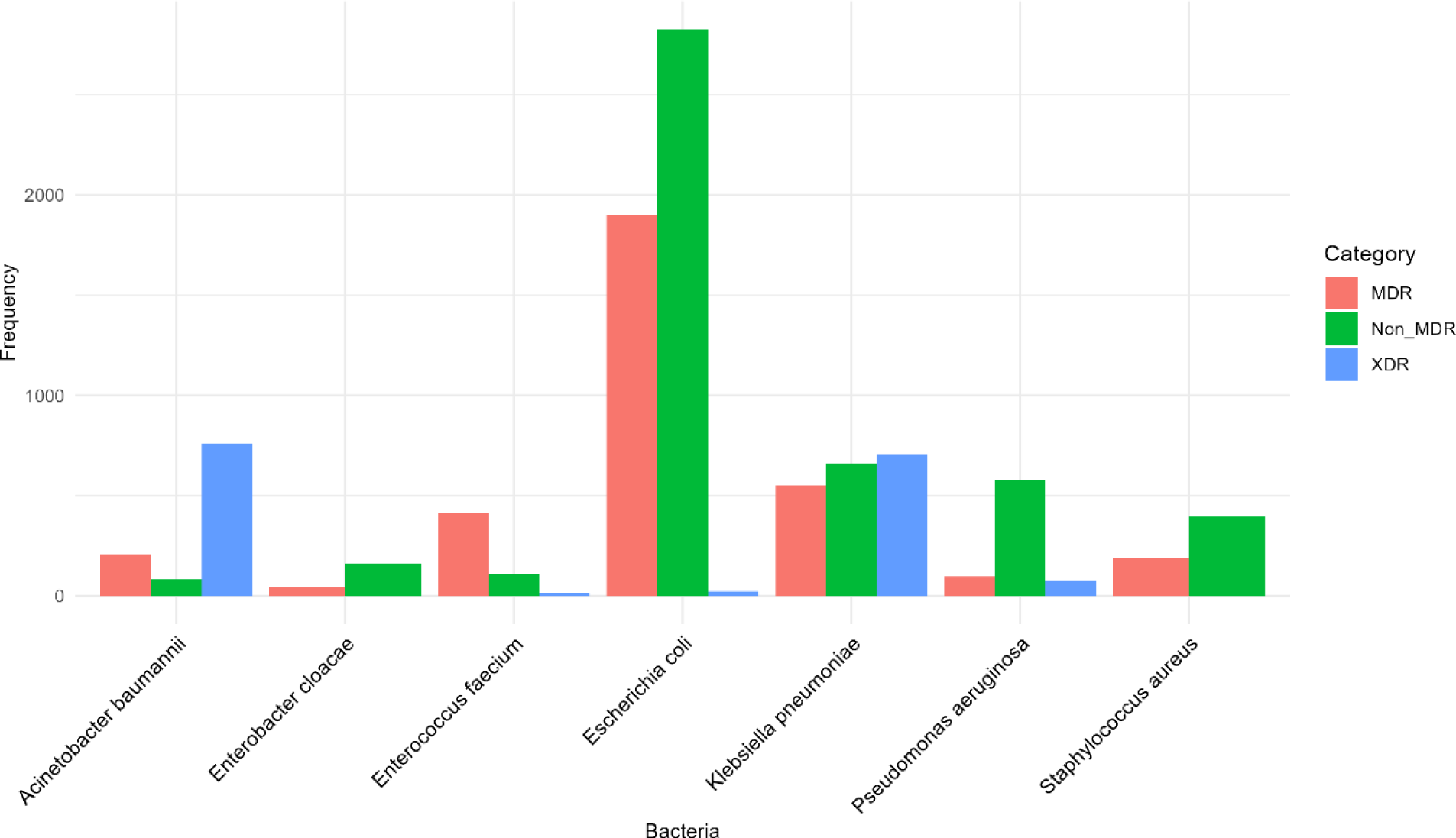
Antibiotics susceptibility category of the ESKAPE bacteria.

### Bacterial to antibiotics

presents the susceptibility of ESKAPE isolates to specific antibiotics for gram-negative and gram-positive bacteria. Among the gram-negative isolates, *A. baumannii* demonstrated high resistance to all the tested antibacterial agents, with high resistance to ciprofloxacin, Piperacillin/tazobactam, imipenem, and amikacin, respectively. In contrast, P. aeruginosa strains were highly susceptible to amikacin, cefepime, ceftazidime, and ciprofloxacin. In addition, clinical success can also be expected with piperacillin-tazobactam empirical treatment since the overall susceptibility rate was 73.9% (**Fig 4**).

Various Klebsiella species have shown varying levels of resistance to specific antibacterial agents. *K. pneumoniae* isolates demonstrated low susceptibility rates to all beta-lactams except for the fourth-generation cephalosporins cefepime and carbapenems (imipenem and ertapenem), indicating extended-spectrum beta-lactamase (ESBL) development. In contrast*, K. oxytoca* strains were susceptible to all the tested antibiotics, with extreme sensitivity to levofloxacin, aminoglycosides, and carbapenems (**Table 4**).

While *E. coli*, was the most often isolated gram-negative bacteria, was sensitive to aminoglycosides and imipenem, accounting for approximately 97.9% of both antibiotics. In contrast, the *E. coli* isolates presented high levels of resistance to ampicillin, Amoxicillin -clavulanic acid, cefixime, ceftazidime cefepime and ciprofloxacin (**Fig4**).

Unfortunately, since all the isolates of *E. cloacae* were resistant to ampicillin, co-amoxiclav, cefixime, and cefoxitin. Interestingly, none of the Enterobacter isolates resisted levofloxacin, amikacin, tobramycin, or imipenem All the isolates tested were sensitive (**Fig 4**). On the other hand, *P. mirabilis* was highly susceptible to beta-lactam agents, ranging between 70% and 97.9%. In contrast, ciprofloxacin susceptibility was 62%, amikacin susceptibility was 97%, tobramycin susceptibility was 83%, and gentamycin susceptibility was 77% (**Table**). ESBL-positive Enterobacterales strains (659/3719; 17.7%) were generally less susceptible to non-beta-lactam antibacterial drugs than were ESBL-negative strains. Unfortunately, these strains are highly resistant to nitrofurantoin, ampicillin, and sulfamethoxazole-trimethoprim.

Interestingly, *M. morganii* was resistant to nitrofurantoin, ampicillin, and coamoxiclav. Fortunately, clinical success can also be expected with ertapenem since the overall susceptibility rate was 95.3%.

Among gram-positive bacteria, *E. faecalis* can be efficiently treated with first-line antibiotics such as ampicillin since most of the isolates tested are sensitive but interestingly show high levels of resistance to sulfamethoxazole-trimethoprim. In contrast, in the case of *E. faecium* isolates, clinical success can be predicted via linezolid, tigecycline, Vancomycin, and Teicoplanin, all preserved antibacterial activity based on in vitro data. *E. faecium* showed high resistance to ampicillin, levofloxacin, and Gentamycin, whereas MDR *E. faecium* was found in two-thirds of the isolates (74.3%) (**Fig 4**).

On the other hand, this study revealed a high frequency of methicillin resistance among coagulase-negative staphylococci, particularly *S. hemolytic* bacteria (98.1%), *S. epidermidis* (88.9%), and *S. hominis* (84%) were detected (**Supplementary 4**).

Moreover, methicillin-resistant *S. aureus* (MRSA) was found in 35.2% of the strains. MRSA did not show a clear increasing or declining trend over time. Methicillin-resistant strains were also far less susceptible to many other antibacterial drugs than methicillin-susceptible strains (MSSA). Interestingly, linezolid, Daptomycin, tigecycline Vancomycin, and Teicoplanin are expected to provide effective MRSA therapies since most isolates tested are susceptible to these drugs. These compounds are also effective therapies for methicillin-resistant coagulase-negative staphylococci (**Table 5**).

**Fig 3.**
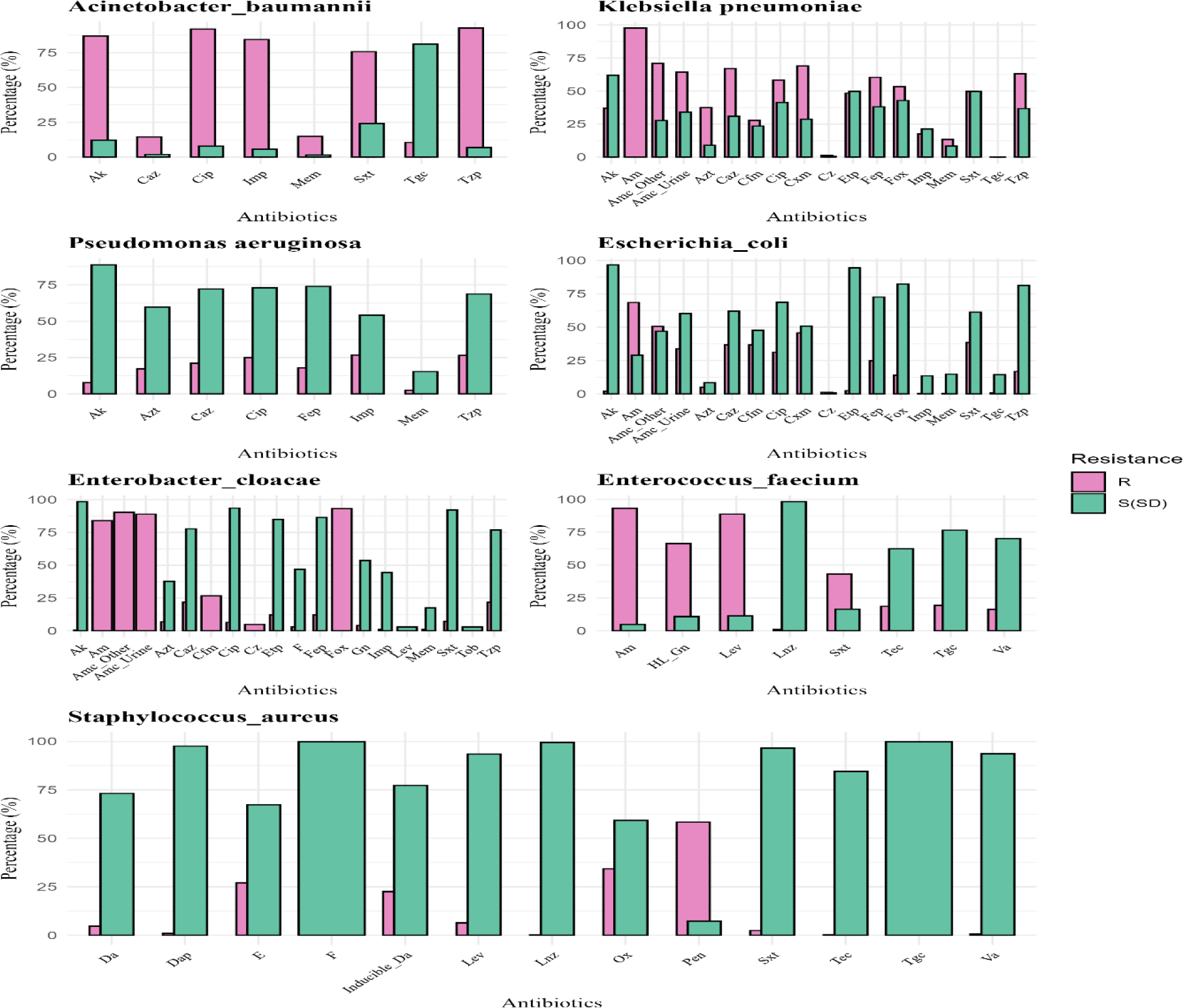
Antibiotic Susceptibility and Resistance Rates Among Major Pathogens AM. Amoxicillin, **AK**: Amikacin, **Tzp**: Piperacillin/tazobactam, **Gm**: Gentamycin, **Azt:** Aztreonam, **Sxt**: Trimethoprim/sulfamethoxazole, **Fep:** Cefepime, **Cip**: Ciprofloxacin, **Caz:** Ceftazidime**, Amc_O:** Amoxicillin + clavulanic acid other**, Cfm:** Cefixime, **Amc_U:** Amoxicillin + clavulanic acid urine, **Etp:** Ertapenem, **Fox:** Cefoxitin, **F:** Nitrofurantoin, **Tgc:** Tigecycline, **Pip:** Piperacillin, **Imp:** Imipenem, **Cxm:** Cefuroxime, **Lev:** Levofloxacin, **Tob:** Tobramycin, **AM:** Amoxicillin, **Ox:** Oxacillin, **DA:** Clindamycin, **E:** Erythromycin, **Sxt:** Sulfamethoxazole, **Lnz:** Linezolid, **Va:** Vancomycin, **Tec:** Teicoplanin, **Tgc:** Tigecycline, **Pen_G**: Penicillin, **Pen:** Penicillin, **Cfx:** Cefoxitin, **HI_Gn:** High Gentamycin, **Lev:** Levofloxacin, **F:** Nitrofurantoin **Ind_Da:** Inducible Clindamycin, **Dap:** Daptomycin

### Variations in Antibiotic Susceptibility across Demographics and Sample Types

The distribution of demographic data shows that the > 60 group was high in MDR and XDR (61.9%), which were remarkable compared to other age groups (Table 6). In gender, results show slight differences between males and females, especially in the XDR group, where males (19.68%) and Females (9.28%) were highly different from females (**Table 6**).

**Table 6:** Variations in Antibiotic Susceptibility across Demographics and Sample Types.

| **Demographics and Sample Types** |  | **NonMDR**<br>**n (%)** | **MDR**<br>**n (%)** | **XDR**<br>**n (%)** |
| --- | --- | --- | --- | --- |
| **Age** | **Pediatric (0-5)** | 1238 (67.7%) | 591 (32.3%) | 1 (1%) |
|  | **School-Age Children (6-17)** | 692 (72.2%) | 260 (27.1%) | 7 (0.7%) |
|  | **Young Adults (18-40)** | 633 (63.2%) | 287 (28.2%) | 82 (8.2%) |
|  | **Middle-Aged Adults (41-60)** | 579 (35.3%) | 451 (45.3%) | 247 (19.3%) |
|  | **Older Adults (61-80)** | 1056 (35.2%) | 1152 (38.4%) | 789 (26.3%) |
|  | **Elderly (80+)** | 616 (35.5%) | 658 (38.0%) | 459 (26.5%) |
| **Gender** | **Female** | 3301 (55.4%) | 1958 (32.8%) | 703 (11.8%) |
|  | **Male** | 1513 (39.4%) | 1441 (37.6%) | 882 (23.0%) |
| **Samples Types** | **Blood** | 274 (31.6 %) | 313 (36.1 %) | 280 (32.3 %) |
|  | **Catheter** | 58 (31.7 %) | 53 (29.0 %) | 72 (39.3%) |
|  | **Rectal Swab** | 25 (19.2 %) | 92 (70.8 %) | 13 (10.0 %) |
|  | **Sputum** | 338 (42.7 %) | 235 (29.7 %) | 219 (27.7 %) |
|  | **Tracheal aspirate** | 145 (18.8%) | 196 (25.4 %) | 431 (70.5%) |
|  | **Urine** | 3788 (56.9%) | 2382 (35.8 %) | 484 (7.3 %) |
|  | **Wound or Abscess** | 185 (46.6%) | 126 (31.7%) | 86 (21.7 %) |
|  | **Stool** | 1 (33.3 %) | 2 (66.7 %) | 0 |

Isolated bacteria from sample types, especially urine, presented a low percentage of MDR and XDR, but they were high-frequency samples. On the other hand, Tracheal aspirate had a low frequency, but the number of MDR and XDR was high (**Table 6**).

### Demographic Analysis

Data reveals significant patterns in antibiotic resistance based on age. Among the children, 32.3% of pediatric and 27.1% of school-age children exhibited MDR characteristics, with only 1% and 0.7 %, respectively, showing XDR properties. On the other hand, among the adult age group, 28.2% of Young Adults, 45.3% of Middle-Aged Adults, and 38.3% of Older Adults categorized as MDR, and 8.2%, 19.3%, and 26.3% showed XDR patterns, respectively. The population over 80 exhibited the highest prevalence XDR (26.5%) pattern, indicating a worrying trend of increasing resistance with age (**Table 6**).

Observations revealed gender disparities in susceptibility patterns. A more significant proportion of non-MDR strains (55.4%) was observed in females, in contrast to males (39.4%). Conversely, males exhibited a higher incidence of MDR (37.6%) and XDR (23%) strains than females MDR (32.8%) and XDR (11.8%). This implies that underlying biological or behavioral factors may influence these outcomes (Table 6). For variations of Antibiotic Susceptibility patterns across different clinical samples, see **SupplementaryFig1**

### Sample Type Analysis

The analysis of various sample types indicates differences in antibiotic susceptibility. Blood samples revealed a significant presence of MDR (36.1 %) and XDR (32.3 %) strains, emphasizing the critical need for effective treatment strategies for bloodstream infections. Catheter samples also exhibited high XDR (72.7 %) rates, suggesting potential transmission and infection control challenges. Urine samples, commonly associated with urinary tract infections, showed a high percentage of non-MDR strains (56.9 %) but also had a proportion of MDR (35.8 %) and a minimal occurrence of XDR (7.3 %). In contrast, sputum samples displayed a higher prevalence of non-MDR (42.7 %), along with significant MDR (29.7 %) and XDR (27.7 %) cases Table 7.

### Distribution of pathogenic bacterial isolates across different hospital department

There were 13378 bacterial isolates of pathogenic bacteria scattered across different hospital department. The Intensive Care Unit (ICU) had the highest number of infections (3261; 33.95%), followed by the pediatric unit (2315;24%). Numerous infections were also reported in patients treated in clinic departments (13.5%), Urology (6.1%), Pulmonary Diseases (4.2%), Gynecology and Obstetrics (3.9%), Emergency (2.9%), and less than 1% in COVID unit, furthermore about 3.16% of pathogenic bacteria isolated from other Partner Hospital.

### Clinical Department Associations

Figure 6 depicts the relationship between clinical departments and susceptibility patterns. The ICU department showed a concerning high prevalence of MDR (35.3%) and XDR (36%) strains. This is followed by the Children’s Diseases department, where 30.3% of isolates are categorized as MDR. The COVID department exhibited approximately an equal distribution of MDR and non-MDR, moreover, Urology, Polyclinic, Emergency, Pulmonary Diseases, and Gynecology and Obstetrics showed a concerning distribution of MDR and these, highlighting the need for ongoing surveillance in emerging infectious diseases (Fig 6).

**Fig 6.**
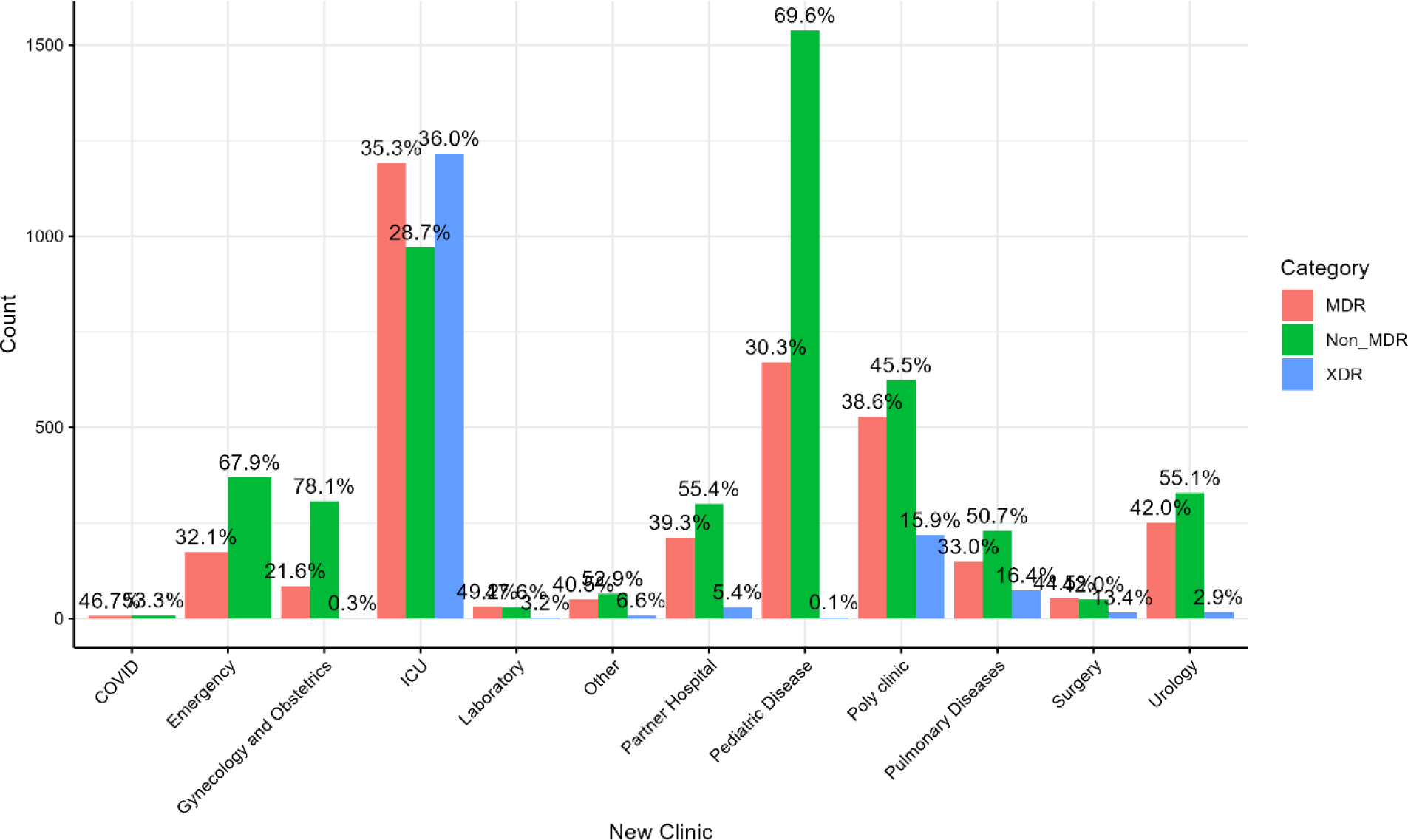
Antibiotic Susceptibility pattern across the Hospital Department.

### Antibiotic Susceptibility across the Hospital Department

Distribution of bacterial resistance across various hospital departments. The resistance to all tested antibiotics varied significantly between hospital units. The hospital units included the pediatric department, intensive care unit, COVID unit, emergency department, gynecology and obstetrics, clinic departments, laboratory department, pulmonary diseases unit, urology department, and surgery unit. Notably, the gynecology and obstetrics clinic was more susceptible than other departments (Table 8).

**Supplementry table 7:** Frequency and precentege of antibiotics in clinical departments.

| Antibiotic | COVID N (R%) | Emergency | Gynecology and Obstetrics | ICU | Laboratory | Other | Partner Hospital | Pediatric Disease | Poly clinic | Pulmonary Diseases | Surgery | Urology |
| --- | --- | --- | --- | --- | --- | --- | --- | --- | --- | --- | --- | --- |
| Amikacin | 0 (0%) | 6 (1.21%) | 2 (0.53%) | 1309 (46.67%) | 4 (6.56%) | 9 (9.89%) | 39 (7.86%) | 23 (1.11%) | 255 (22.81%) | 88 (22.51%) | 17 (19.32%) | 26 (4.79%) |
| Amoxicillin | 9 (81.82%) | 350 (72.31%) | 206 (56.44%) | 1788 (91.18%) | 43 (89.58%) | 69 (85.19%) | 364 (79.48%) | 1517 (75.25%) | 766 (79.87%) | 231 (95.45%) | 68 (94.44%) | 405 (77%) |
| Amoxicillin-Clavulanate (Other) | 6 (66.67%) | 238 (50.42%) | 121 (33.15%) | 1247 (74.14%) | 29 (60.42%) | 49 (59.76%) | 247 (54.89%) | 1036 (52.83%) | 515 (59.95%) | 158 (73.83%) | 41 (65.08%) | 268 (53.07%) |
| Amoxicillin-Clavulanate (Urine) | 3 (37.5%) | 173 (37.86%) | 72 (20.06%) | 1064 (64.29%) | 18 (39.13%) | 35 (43.21%) | 174 (40%) | 776 (40.71%) | 380 (45.45%) | 124 (59.9%) | 32 (52.46%) | 177 (35.83%) |
| Aztreonam | 2 (50%) | 14 (29.79%) | 5 (13.51%) | 766 (62.74%) | 5 (33.33%) | 12 (54.55%) | 12 (30%) | 25 (17.48%) | 159 (46.09%) | 67 (30.32%) | 17 (34%) | 15 (31.91%) |
| Ceftazidime | 4 (36.36%) | 168 (33.87%) | 71 (19.19%) | 1350 (62.91%) | 24 (40.68%) | 48 (53.33%) | 217 (44.11%) | 668 (32.47%) | 445 (44.95%) | 131 (41.19%) | 48 (60.76%) | 215 (39.74%) |
| Cefoxitin | 0 (0%) | 13 (46.43%) | 6 (42.86%) | 99 (41.42%) | 0 (0%) | 13 (61.9%) | 12 (42.86%) | 22 (40%) | 55 (40.44%) | 7 (31.82%) | 7 (33.33%) | 3 (27.27%) |
| Cefixime | 2 (40%) | 192 (43.54%) | 74 (22.49%) | 435 (60.58%) | 22 (59.46%) | 41 (61.19%) | 225 (53.19%) | 746 (40.63%) | 290 (48.82%) | 70 (73.68%) | 16 (72.73%) | 223 (46.85%) |
| Ciprofloxacin | 5 (45.45%) | 114 (22.66%) | 56 (14.85%) | 1957 (69.01%) | 31 (50%) | 46 (46.94%) | 207 (41.24%) | 292 (13.98%) | 588 (52.08%) | 201 (51.15%) | 48 (53.93%) | 220 (39.71%) |
| Cefuroxime | 5 (55.56%) | 202 (43.63%) | 90 (25.14%) | 1213 (74.92%) | 28 (59.57%) | 50 (63.29%) | 241 (54.52%) | 795 (41.86%) | 460 (56.37%) | 132 (68.39%) | 45 (75%) | 239 (49.9%) |
| Cefazolin | 1 (100%) | 2 (66.67%) | 2 (28.57%) | 40 (72.73%) | 2 (40%) | 0 (NA%) | 4 (100%) | 12 (44.44%) | 12 (70.59%) | 2 (66.67%) | 4 (100%) | 6 (75%) |
| Erythromycin | 1 (50%) | 9 (33.33%) | 5 (35.71%) | 59 (26.34%) | 0 (0%) | 5 (23.81%) | 8 (29.63%) | 17 (30.91%) | 38 (30.16%) | 4 (19.05%) | 8 (42.11%) | 3 (27.27%) |
| Ertapenem | 0 (0%) | 7 (1.49%) | 6 (1.65%) | 743 (44.25%) | 3 (6.25%) | 8 (9.76%) | 36 (8.07%) | 49 (2.51%) | 145 (16.88%) | 38 (17.59%) | 17 (26.98%) | 17 (3.37%) |
| Nitrofurantoin | 0 (0%) | 8 (1.69%) | 2 (0.58%) | 165 (17.03%) | 2 (5.13%) | 6 (6.82%) | 35 (7.64%) | 25 (1.29%) | 61 (8.25%) | 11 (9.32%) | 3 (6.98%) | 35 (7.11%) |
| Fusidic Acid | 0 (0%) | 2 (7.14%) | 1 (7.14%) | 14 (5.83%) | 0 (0%) | 0 (0%) | 1 (3.57%) | 5 (9.09%) | 9 (6.62%) | 1 (4.35%) | 3 (14.29%) | 2 (18.18%) |
| Cefepime | 4 (40%) | 93 (18.98%) | 41 (11.2%) | 1098 (54.71%) | 18 (31.03%) | 31 (34.83%) | 172 (34.96%) | 403 (19.98%) | 340 (35.56%) | 118 (36.88%) | 33 (45.83%) | 148 (27.82%) |
| Cefoxitin | 0 (0%) | 86 (18.42%) | 27 (7.5%) | 878 (52.8%) | 6 (12.77%) | 21 (26.25%) | 80 (18.31%) | 377 (19.54%) | 238 (28%) | 76 (35.85%) | 20 (33.33%) | 76 (15.17%) |
| Gentamicin | 0 (0%) | 48 (10.64%) | 27 (7.89%) | 364 (36.77%) | 8 (16%) | 12 (17.65%) | 70 (15.7%) | 169 (8.64%) | 163 (24.04%) | 33 (28.95%) | 16 (47.06%) | 81 (15.73%) |
| High-Level Gentamicin | 0 (NA%) | 7 (100%) | 0 (0%) | 207 (89.61%) | 0 (NA%) | 1 (100%) | 6 (85.71%) | 34 (77.27%) | 69 (87.34%) | 17 (85%) | 5 (71.43%) | 13 (65%) |
| Imipenem | 1 (25%) | 1 (2.08%) | 0 (0%) | 1137 (60.8%) | 2 (11.76%) | 9 (31.03%) | 5 (12.5%) | 2 (1.26%) | 181 (40.58%) | 78 (27.18%) | 15 (26.79%) | 6 (10.91%) |
| Inducible Clindamycin | 1 (50%) | 7 (25%) | 3 (21.43%) | 51 (21.25%) | 0 (0%) | 5 (23.81%) | 8 (28.57%) | 12 (21.82%) | 31 (22.79%) | 4 (17.39%) | 6 (28.57%) | 3 (27.27%) |
| Levofloxacin | 1 (20%) | 15 (36.59%) | 4 (19.05%) | 426 (58.44%) | 1 (14.29%) | 1 (4.17%) | 11 (25.58%) | 51 (30%) | 131 (45.8%) | 33 (48.53%) | 16 (41.03%) | 26 (43.33%) |
| Meropenem | 0 (NA%) | 0 (0%) | 0 (0%) | 331 (62.34%) | 0 (NA%) | 6 (13.95%) | 14 (9.86%) | 0 (0%) | 66 (33%) | 19 (22.35%) | 2 (14.29%) | 3 (4.05%) |
| Oxacillin | 0 (0%) | 12 (44.44%) | 5 (38.46%) | 79 (35.75%) | 0 (0%) | 9 (52.94%) | 12 (42.86%) | 20 (37.04%) | 47 (36.72%) | 6 (28.57%) | 6 (30%) | 3 (27.27%) |
| Penicillin | 2 (100%) | 16 (94.12%) | 10 (83.33%) | 149 (89.76%) | 0 (0%) | 0 (NA%) | 4 (44.44%) | 35 (92.11%) | 87 (87.88%) | 11 (100%) | 19 (95%) | 6 (100%) |
| Piperacillin | 0 (0%) | 1 (50%) | 5 (83.33%) | 41 (56.16%) | 3 (50%) | 0 (0%) | 3 (50%) | 26 (57.78%) | 14 (53.85%) | 5 (50%) | 1 (25%) | 10 (47.62%) |
| Trimethoprim-Sulfamethoxazole | 6 (54.55%) | 130 (25.34%) | 92 (23.71%) | 1763 (60.52%) | 28 (49.12%) | 34 (30.36%) | 201 (40.36%) | 680 (32.18%) | 515 (42.63%) | 148 (45.54%) | 41 (40.59%) | 203 (36.45%) |
| Tobramycin | 0 (0%) | 1 (50%) | 1 (16.67%) | 119 (58.91%) | 0 (0%) | 0 (0%) | 1 (16.67%) | 4 (8.33%) | 26 (53.06%) | 5 (27.78%) | 4 (50%) | 6 (27.27%) |
| Tobramycin | 1 (9.09%) | 83 (16.77%) | 47 (12.5%) | 1794 (63.68%) | 11 (18.03%) | 25 (25.51%) | 118 (24.13%) | 398 (19.42%) | 433 (38.97%) | 156 (39.49%) | 38 (43.18%) | 117 (21.43%) |
| Vancomycin | 0 (0%) | 1 (2.63%) | 0 (0%) | 67 (14.14%) | 0 (0%) | 0 (0%) | 1 (3.57%) | 5 (4.5%) | 14 (6.57%) | 1 (2.27%) | 0 (0%) | 3 (8.11%) |

## Discussion

With an emphasis on the therapeutic significance of these microbes, the global concern in managing infectious diseases has grown due to the evolution of antibiotic resistance in ESKAPE pathogens [15].

This study was aimed to investigate the bacterial spectra and susceptibility patterns of ESKAPE pathogens isolated from different clinical departments at Nigde Omer Halisdemir University Education and Research Hospital in Nigde, Turkey. The findings of this study draw attention to the concerning patterns of resistance that have emerged among ESKAPE bacteria in Turkish hospitalized patients, which the WHO has classified as critically important and a top priority for developing new drugs. The work closes a substantial gap in knowledge. Furthermore, epidemiological data from other clinical departments are limited globally. Most of these studies focus on specific diseases or specimen types or primarily on certain bacteria or bacterial families, making meaningful benchmarking impossible. However, at this time, we refer to the most recent report from the European Surveillance of Antimicrobial Resistance Network (EARS-NET), which compares data on antibiotic resistance for specific bacterial illnesses across Member States and offers current resistance levels [16, 17]. According to this study, ESKAPE pathogens were the most prevalent bacterial isolated strains, responsible for 73.2% (9798 /13378) of all the clinical isolates. We detected a predominance of gram-negative bacteria, and the most common isolates were from the Enterobacterales family. Within this group, *E. coli* 4747 (35.5%) was the most frequently isolated organism followed by *K. pneumonia* 1921 (14.3%) and *A. baumannii* 1049 (7.8%) with an alarming XDR pattern among *A. baumannii* which exceeding 72.3% to most antimicrobial agents, including 1^st^,2^nd^,3^rd^ and 4^th^ generation of cephalosporins, carbapenems, aminoglycosides and fluoroquinolones. On the other hand, our study showed that the most common clinical samples were midstream urine \ urinary catheter samples, followed by blood culture samples and Tracheal aspirate. In regard to the isolates from urinary tract infections (UTIs), *E. coli* and *K. pneumoniae* were documented as major bacterial agent which is consistent with a study conducted from Germany, which revealed that the Enterobacterales group was the most commonly isolated pathogen from urine sample and blood cultures, followed by *S. aureus*. A study from the USA also demonstrated that *E. coli* and *Klebsiella.spp* were the most frequently isolated bacteria from UTIs [18, 19]. Compared with studies focusing on ESKAPE pathogens reported from Hungary, ESKAPE pathogens account for 72.22% of the total isolates, which is approximately same as noted in our study [20]. In the same study*, E. coli* was the most common isolate (44.1%), followed by the Klebsiella genus 13. 4%. The third most common isolate was *S. aureus* (11.3%), which was higher than that reported in our study [21]. Other specimens, such as wound swabs, sputum, and discharges, revealed S. aureus and *K. pneumoniae* to be significant infectious agents. Compiling such a laboratory database can provide vital insight into the different types of samples and came across drug resistance isolates in regions where microbiological investigations are scarce. As a result, our findings can be utilized to support existing empirical therapies [22]. On the other hand, this study also evaluated variations in antimicrobial resistance for ESKAPE pathogens according to the patient age group (Pediatric, School-Age Children, Young Adults, Middle-Aged Adults, Older Adults and Elderly). Overall, we find that isolates from older patients had greater levels of antibiotic resistance than isolates from children and adults, with MDR and XDR being more common in the elderly group at 45% and 16.9%, respectively. In comparison, children account for 30.09% and 0.04%, respectively. Furthermore, in this study, the intensive care unit (ICU) and pediatrics department had the highest frequency of bacterial infections, indicating that babies and the elderly may be more vulnerable to bacterial infections. As a result, adequate attention needs to be given, particularly to preserving proper hygiene in the ICU [23]. We isolated *P. aeruginosa, E. cloacae, E. faecalis, E. faecium, P. mirabilis*, and *S. pneumoniae* from patients in various clinical departments; this occurrence is alarming, as these bacteria are primarily regarded as hospital-acquired pathogens and are frequently linked to infections in immunocompromised patients. Currently, MDR-XDR-PDR bacterial infections caused by ESKAPE organisms represent a significant health issue and substantially burden effective therapeutic options. These classifications are extensively employed and acknowledged for their relevance in microbiology and epidemiology [13]. This study classified 34.7 % of the isolates as MDR, whereas 16.2 % were XDR. This percentage was higher than that reported in Italy, with an MDR of 10.67% [24], and Hungary, with an MDR of 23.8% [20]. MDR was found in a broad spectrum of ESKAPE isolates, with *E. faecium* being the most common, followed by *S. aureus, K. pneumoniae, A. baumannii, E. cloacae*, and *P. aeruginosa*. Moreover, *A. baumannii* had the highest prevalence of XDR, followed by *K. pneumoniae*, *P. aeruginosa*, and *E. faecium*, with the lowest prevalence of *E. coli* and *P. mirabilis*. MDR was detected among Enterobacterales bacteria, and the highest prevalence rates of MDR were found in *P. mirabilis* (48.2%), *E. coli* (40%), *Klebsiella spp*. (29.9%) *and E. cloacaceae* (20.1%). This number was higher than that reported in the Hungarian study, which reported *Proteus* spp. MDR rates of 40.68%, *Klebsiella spp*. of 27.41%, *E. coli* 26.07%, *and Enterobacter spp.* 11.76% [20]. In contrast to nonfermenting gram-negative bacteria, this study revealed lower MDR rates than a study reported in Hungary showed that the MDR rate of *P. aeruginosa* was 13.26% (vs 13.1% in our study), whereas that of *A. baumannii* was 13.26%. 67.67% (vs. 22% in our study), similar to a Korean multicenter study [25]. Moreover, the highest prevalence rates of XDR among gram-negative bacteria were found in *A. baumannii* (69.1%) and *K. pneumoniae* (27.4%), and no XDR was reported in *E. cloacae*. Compared with the Hungarian study, no XDR was noted among all the isolates except for one isolate of *P. aeruginosa* [20]. Variations in carbapenem resistance trends exist within Arab League countries and throughout Europe. *The* prevalence of carbapenem-resistant *A. baumannii* isolates in northern and eastern Europe and the Arab League’s Levant region, which includes Syria, Jordan, Lebanon, Palestine, and Iraq, has increased [26, 27].

*A. baumannii* demonstrated high resistance to all the tested antibacterial agents and extreme resistance to all the tested beta-lactams, with a high level of resistance to imipenem (92.5%), the same as that reported in the study in Turkey by Tuna [28], which indicates that carbapenem-resistant *A. baumannii* (CRAB) and no marked decrease in resistance to b-lactams are observed when a b-lactamase inhibitor such as tazobactam is used. In contrast, *P. aeruginosa* strains were highly susceptible to cefepime (82.7%), ceftazidime (78.7%), and amikacin (92.5%). In addition, clinical success can also be expected with piperacillin-tazobactam empirical treatment since the overall susceptibility rate was 73.9%. Various Klebsiella species have shown varying levels of resistance to specific antibacterial agents. *K. pneumoniae* isolates presented low susceptibility rates to all beta-lactam generations except the fourth-generation cephalosporins cefepime and carbapenems (imipenem and ertapenem); on the other hand, resistance to beta-lactam and beta-lactamase inhibitors such as co-amoxiclav and piperacillin-tazobactam confirmed ESBL production by these strains, with MDR (31.6%) and XDR (27.4%).

In contrast, the *K. oxytoca* strains were susceptible to all the tested antibiotics, with extreme sensitivity to levofloxacin, aminoglycosides, and carbapenems. *E. coli*, the most often isolated gram-negative bacteria, was sensitive to aminoglycosides and imipenem, accounting for approximately 97.9% of both antibiotics.

In contrast, the *E. coli* isolates presented high levels of resistance to cefixime, ampicillin, piperacillin, cefepime, and coamoxiclav. Unfortunately, since all the isolates tested were resistant, E. cloacae were resistant to ampicillin, co-amoxiclav, cefixime, and cefoxitin. Interestingly, the Enterobacterales isolates’ resistance patterns differed from those reported in our study. Research conducted at Karabük University Training and Research Hospital in Turkey from January 2016 to December 2020 revealed that *E. cloacae* was prevalent in 59.2% of clinical samples from outpatients and inpatients. Among these samples, 29% demonstrated resistance to ceftazidime. In contrast, the present study revealed a lower prevalence of *E. cloacae*, at 1.6% in all bacteria and 2.5% in ESKAPE bacteria, with 21.7% of the cases demonstrating resistance to ceftazidime. However, the imipenem resistance percentages in this study were lower than those reported in the Turkish study, which reported a 3% resistance rate [29]. On the other hand, *P. mirabilis* was highly susceptible to beta-lactam agents, ranging between 70% and 97.9%. Ciprofloxacin susceptibility was 62%, amikacin susceptibility was 97%, tobramycin susceptibility was 83%, and gentamycin susceptibility was 77% (Table 4). Additionally, 48.3% of the samples were from children’s diseases, and 19.4% were from the ICU. ESBL-positive Enterobacterales strains (659/3719; 17.7%) were generally less susceptible to non-beta-lactam antibacterial drugs than were ESBL-negative strains. Unfortunately, it is highly resistant to nitrofurantoin, ampicillin, and sulfamethoxazole-trimethoprim. Interestingly, *M. morganii* was resistant to nitrofurantoin, ampicillin, and coamoxiclav. Fortunately, clinical success can also be expected with ertapenem since the overall susceptibility rate was 95.3%.

MDR bacteria, on the other hand, were found in gram-positive bacteria. MDR prevalence rates were highest for *S. hemolyticus* (97.5%), *S. epidermidis* (85.5%), *S. hominis* (84.4%), *E. faecium* (74.3%), *S. pneumonia* (41%), and *S. aureus* (35.7%). *E. faecium* had the highest XDR prevalence rate among gram-positive bacteria (3.6%), and no XDR was discovered in any of the other isolated gram-positive species. Among the S. aureus isolates, 35.2% were identified as methicillin-resistant strains (MRSA). MRSA is also considerably less susceptible to many other antibiotics than methicillin-susceptible strains (MSSA). This percentage was more significant than that reported by Benkő, Ria et al. from Hungary, in which MRSA was isolated in 16.9% of the population [20], and lower than that reported in Saudi Arabia, in which MRSA was isolated in 55.3% of the population [30]. On the other hand, this study revealed a high frequency of methicillin resistance among coagulase-negative staphylococci, particularly *S. hemolyticus* (98.1%), *S. epidermidis* (88.9%) and *S. hominis* (84%). The results of this study were more significant than those reported in Germany, where methicillin resistance was detected among coagulase-negative *S. hemolyticus* and *S. epidermidis* at 7% [31]. Interestingly, linezolid and tigecycline are expected to provide effective MRSA therapies since all the isolates tested are susceptible to these drugs. They are also effective therapies for methicillin-resistant coagulase-negative staphylococci. Concerning *Enterococcus spp.*, 99.14% of the vancomycin-susceptible *E. faecalis* isolates were resistant (resistance rate less than 1%), whereas 72.8% of the vancomycin-susceptible *E. faecium* isolates were resistant (resistance rate 27.2%). This rate is much higher than the EU mean resistance of *E. faecium* to vancomycin (17.3%) and similar to the value reported in 2018. In contrast, it was 10.4% (95% CI 10–11) in 2014 in countries of the European Union and European Economic Area (EU/EEA)[32]. In a hospital-based study conducted in Italy, among 1628 isolates from community-acquired infections, 19 were identified as *E. faecium*, with vancomycin resistance detected in only 1 case, corresponding to 5.3% [33]. A recent German study reported no isolation of VRE [19]. These findings indicate that if the pathogenic role of *E. faecium* is suspected, one should consider the possibility of glycopeptide resistance. Fortunately, clinical success can be predicted using linezolid and tigecycline, both useful antibacterial drugs, based on in vitro data.

In this study, *E. coli* and *K. pneumoniae* were identified as the leading bacterial agents in the isolates from the urology department, which aligns with recent investigations carried out in Ethiopia, Gabon, and Egypt [34–37]. According to this study, CoNS and *K. pneumonia* were the primary bacterial isolates from bloodstream infection, especially among the elderly and children, which is nearly similar to many studies [38, 39]. According to this study, the primary bacterial isolate in bloodstream infection is CoNS, followed by K. pneumoniae, and mainly isolated from the ICU and pediatric unit, which are considered the major critical illness clinical departments. According to this finding, CoNS was the primary pathogen in bloodstream infection. The significant incidence of CoNS could be attributed to poor sanitary care and rising nosocomial infections in our hospitals. A recent survey shows that hospital-acquired bacterial illnesses are more common in poor nations [38]. On the other hand, *A. baumannii, S. aureus, E. coli*, and *K. pneumoniae* were found to be primary infectious agents among the other specimens like tracheal aspirate, wound, and sputum, respectively. Among ESKAPE pathogens*, E. coli, A. baumannii,* and *K. pneumoniae* were the most common infectious agents isolated from the ICU. Also, *E. coli* was reported as a common bacterial isolated from the gynecology and obstetrics department. It has been connected to endometriosis, intrauterine growth restriction, low birth weight, preterm delivery, and preeclampsia[40, 41]. Additionally, our study revealed that during the COVID-19 pandemic peak, there was a noticeable increase in the infection of *K. pneumoniae* and *E. coli*.

The study found notable differences in resistance levels to several antibiotics among hospital departments, including Amikacin, Gentamycin, Tobramycin, Cefepime, Ceftazidime, Cefuroxime, Cefixime, Cefoxitin, Imipenem, Tigecycline, Ciprofloxacin, Levofloxacin, Vancomycin, and Linezolid (Table 8). Since multiple strains of the same microbe may cause an infection, these findings could have implications for empirical therapy for various infectious diseases. Generally, resistance levels of aminoglycosides (Amikacin, Gentamycin, and Tobramycin), fluoroquinolones (Ciprofloxacin and Levofloxacin), Vancomycin, and Linezolid were higher in the ICU compared to other hospital departments. This may indicate the distribution of MDR and XDR strains in the ICU. On the other hand, the resistance level of 3rd generation cephalosporin (Ceftazidime, Cefuroxime, and Cefixime) was also higher in the ICU, followed by the pulmonary disease, surgery unit, urology, and pediatric department compared to the other departments. This finding confirmed the distribution of extended B-lactamase strains across these departments. Cefepime and Imipenem resistance was also higher across ICUs, confirming the distribution of Carbapenemase strains in these departments. The growing trends and long-term high resistance have implications for empirical therapy of infectious disease across different hospital departments, Turkey’s most common method of treating bacterial illness. Antimicrobial stewardship implementation strategies are urgently needed to reverse the observed trends, which would be more effective if tailored to specific scenarios or antimicrobials [22, 42].

On the other hand, our findings teach us a vital infection control lesson: department-specific surveillance and monitoring, which play a crucial role in countering antibiotic resistance. The differences in susceptibility patterns amongst clinical departments underscore the need for infection control methods adapted to departmental data. In the context of the results provided in Table 7, our data shows a significant variation in susceptibility patterns across clinical departments. For example, the ICU department had a worrisome number of XDR strains. Whereas the pediatric unit had a more significant rate of non-MDR strains. This highlights the importance of focused actions and surveillance measures tailored to each department’s risks and problems. Moreover, the high incidence of XDR strains in ICUs serves as a significant warning indication that proactive and ongoing surveillance is required in high-risk regions where multidrug-resistant pathogens are more common. This underlines the need for solid infection control techniques, antimicrobial stewardship programs, and monitoring systems to avoid the spread of highly resistant organisms. Interestingly, our study also shows the equal distribution of MDR and non-MDR patients in the COVID-19 unit, emphasizing the unexpected nature of emerging infectious illnesses and the importance of adopting rapid response and surveillance methods. These findings highlight the importance of heightened attention, early detection, and effective containment techniques in dealing with new and emerging infectious dangers. Finally, the disparities in susceptibility patterns between clinical departments highlight the need for interdisciplinary collaboration among physicians, microbiologists, infection control practitioners, and other healthcare workers. A multidisciplinary strategy that blends clinical expertise with microbiological data is critical for developing targeted therapies, improving antibiotic use, and reducing the spread of antimicrobial resistance.

## Conclusions

This research presents significant discoveries about the spread, frequency, and resistance of ESKAPE pathogens in medical environments to antibiotics. The study focused primarily on elderly individuals, with a Fig 1 proportion of female participants. ESKAPE pathogens constitute most bacterial isolates, particularly gram-negative bacteria, including *E. coli*. This study highlights the troubling levels of MDR and XDR strains among these pathogens, especially *A. Baumann and S. hemolytic*.

The data provide a crucial understanding of the resistance patterns of these bacteria across different demographics and clinical specimens, revealing a higher prevalence of resistance in older patients and specific sample types. These findings stress the importance of targeted monitoring and tailored antibiotic stewardship programs in hospital settings to counter the escalating threat of antibiotic-resistant infections. Furthermore, further investigations are needed to elucidate the mechanisms contributing to these resistance patterns and to devise effective strategies for managing infections caused by ESKAPE pathogens. The results underscore the pressing need for improved diagnostic initiatives and treatment guidelines to enhance patient outcomes in light of increasing antibiotic resistance.

The high rates of resistance necessitate the implementation of robust antibiotic stewardship programs to mitigate the spread of resistant strains and improve patient outcomes. Additionally, this study highlights the importance of ongoing research to monitor resistance patterns and inform treatment guidelines, particularly in low—and middle-income countries where the burden of antibiotic resistance is increasing.

The growing presence of MDR and XDR strains, particularly among older demographics and specific clinical samples, requires a comprehensive approach to infection control and the careful use of antibiotics. Continual monitoring and customized treatment strategies are crucial in effectively addressing the increasing threat of antimicrobial resistance. Future research should prioritize understanding the underlying mechanisms of resistance and developing specific interventions to reduce its impact on diverse patient populations.

Our findings highlight the importance of department-specific surveillance, constant monitoring of susceptibility patterns, and proactive infection control techniques adapted to each clinical department’s unique characteristics. Understanding and managing these variances enables healthcare facilities to strengthen infection control methods, improve patient outcomes, and successfully battle antibiotic resistance.

## Data Availability

All data produced in the present study are available upon reasonable request to the authors

## Availability of data and materials

According to Turkey Data Privacy and the Non-Interventional Clinical Research Ethics Committee at Nigde Omer Halisdemir University, the patient data utilized in this study are private. Patient-related information could be used to identify the patient, such as the date of ward admission, age, sex, underlying illness, and antimicrobial profile. The Data Privacy Act permits the acquisition of the study’s data and condensed form to preserve participant and patient privacy. In order to acquire access to raw data, interested researchers who meet the requirements may contact one of the corresponding authors (e.g.). This request has been accepted by the Nigde Omer Halisdemir University Ethics Committee for Non-Interventional Clinical Research.

## Acknowledgments

I want to express my deep appreciation to the director of Nigde Ömer Halisdemir University Training and Research Hospital for their invaluable support and for permitting us to conduct this research. Furthermore, I extend my heartfelt thanks to the Central Lab, notably the Microbiology Lab, and all the dedicated staff in the Microbiology Laboratory., and grateful for Dean of the faculty of science, and all the Faculty of Sciences staff, especially the Biotechnology Department.

## Authors’ contributions

All authors contributed to the manuscript according to the recommendations of the International Committee of Medical Journal Editors. MAS, SBD, and AOT were responsible for designing the study, conducting statistical calculations, interpreting the data, and drafting the manuscript. RK was involved in collecting the raw data. MAS, NMH, and FP contributed to statistical calculations. All authors have read and agreed to the final draft before submission.

## Consent for publication

Not applicable.

## Abbreviations

AMR: Antimicrobial resistance
CLSI: Clinical and laboratory standards institute
ESKAPE: *Enterococcus faecium, Staphylococcus aureus, K. pneumoniae, Acinetobacter baumannii, Pseudomonas Aeruginosa, and Enterobacter spp*
CoNS: Coagulases Negative Staphylococci
AST: Antimicrobial susceptibility testing
EUCAST: The European Committee on Antimicrobial Susceptibility Testing
LIS: Laboratory Information System
MDR: multidrug-resistant
XDR: Extensively drug-resistant
PDR: Pan drug-resistant
WHO: World Health Organization’s
ICU: intensive care unit
ESBL: extended-spectrum beta-lactamase,
MRSA: methicillin-resistant *Staphylococcus aureus*
MSSA: methicillin-susceptible strains,

---

**Supplementry fig1.**
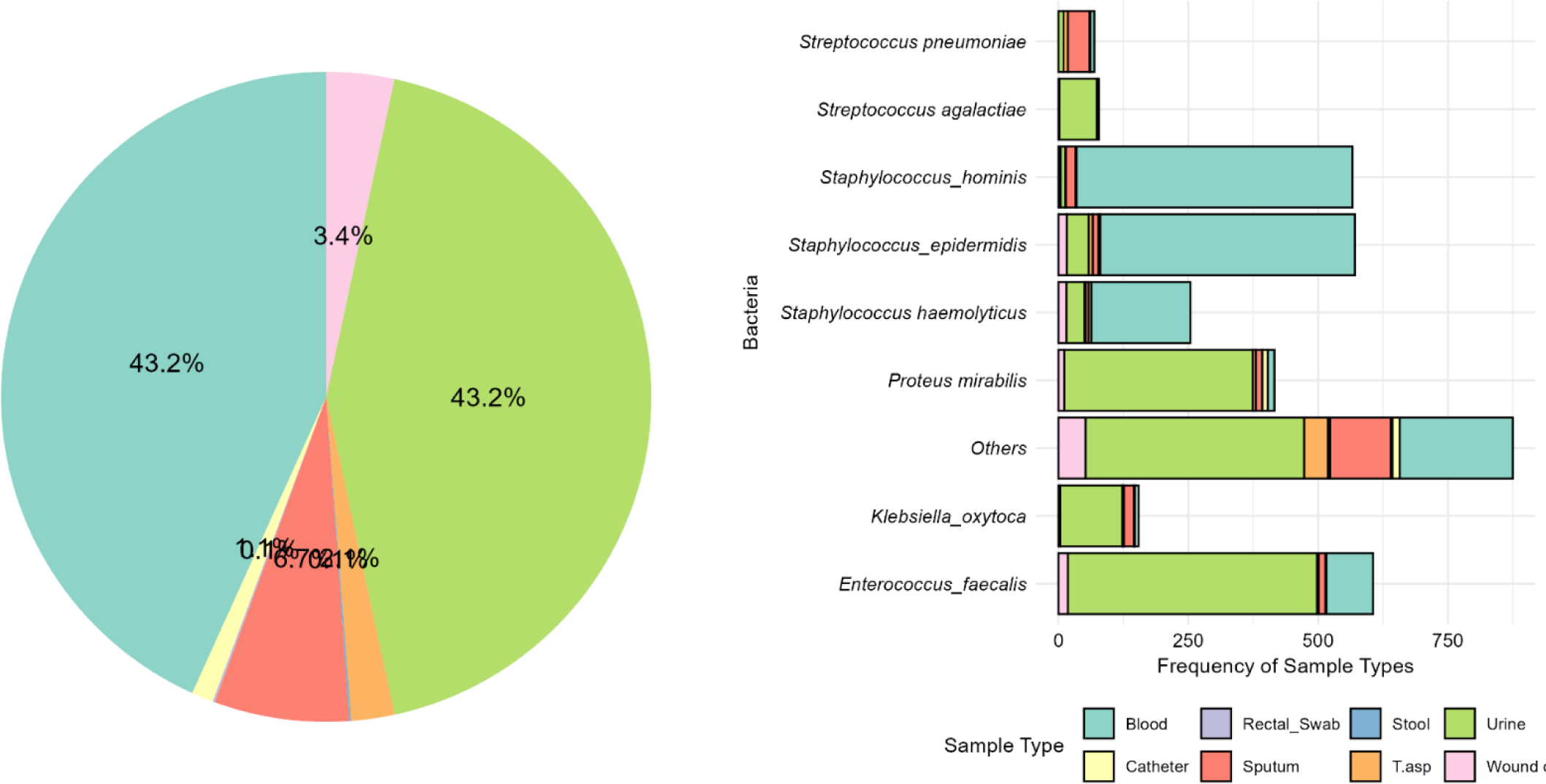
Distribution of bacteria isolated from clinical samples.

**Supplementary Fig 2:**
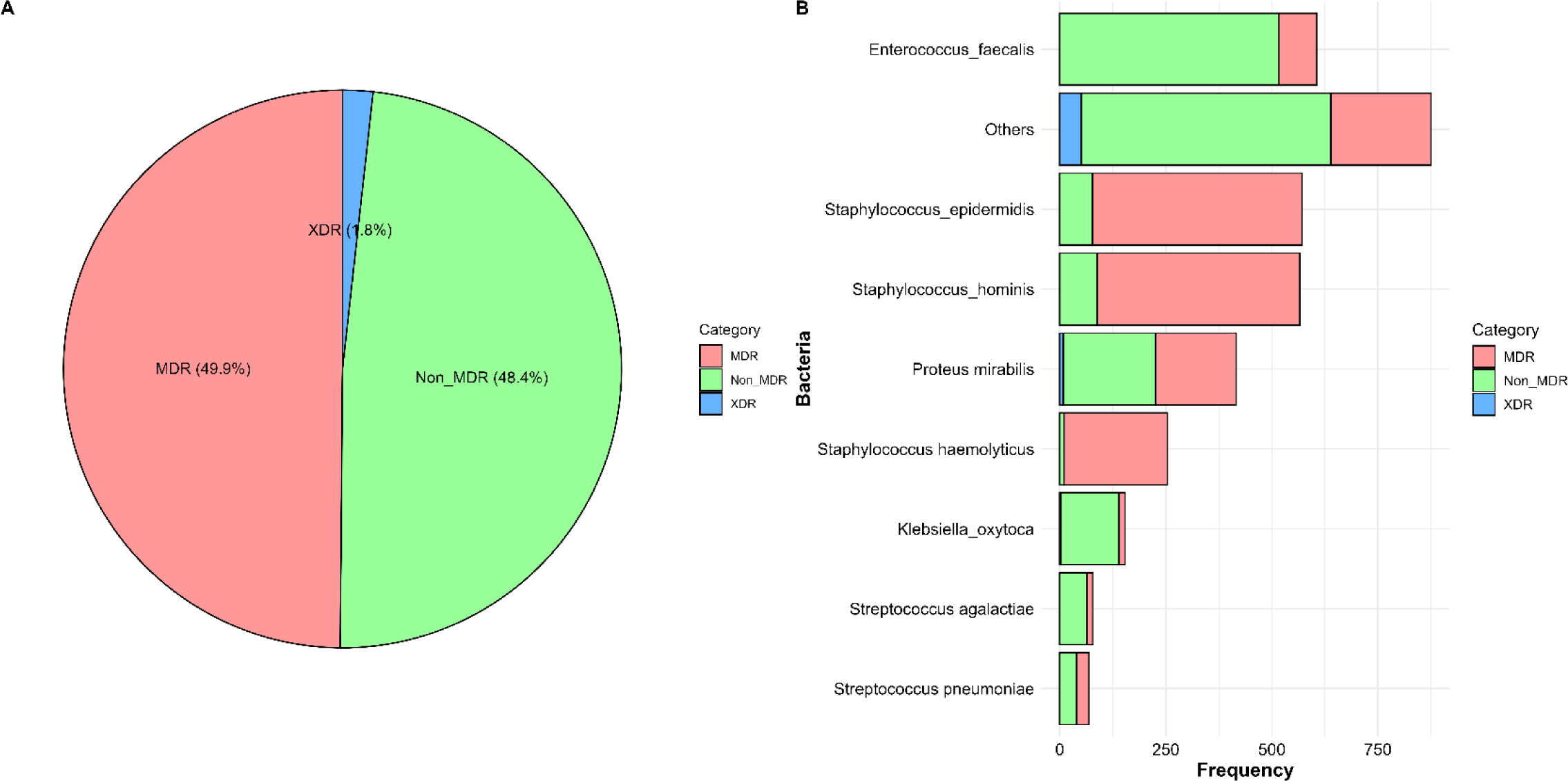
Antibiotic Susceptibility Categories of Multidrug-Resistant Isolated Bacteria.

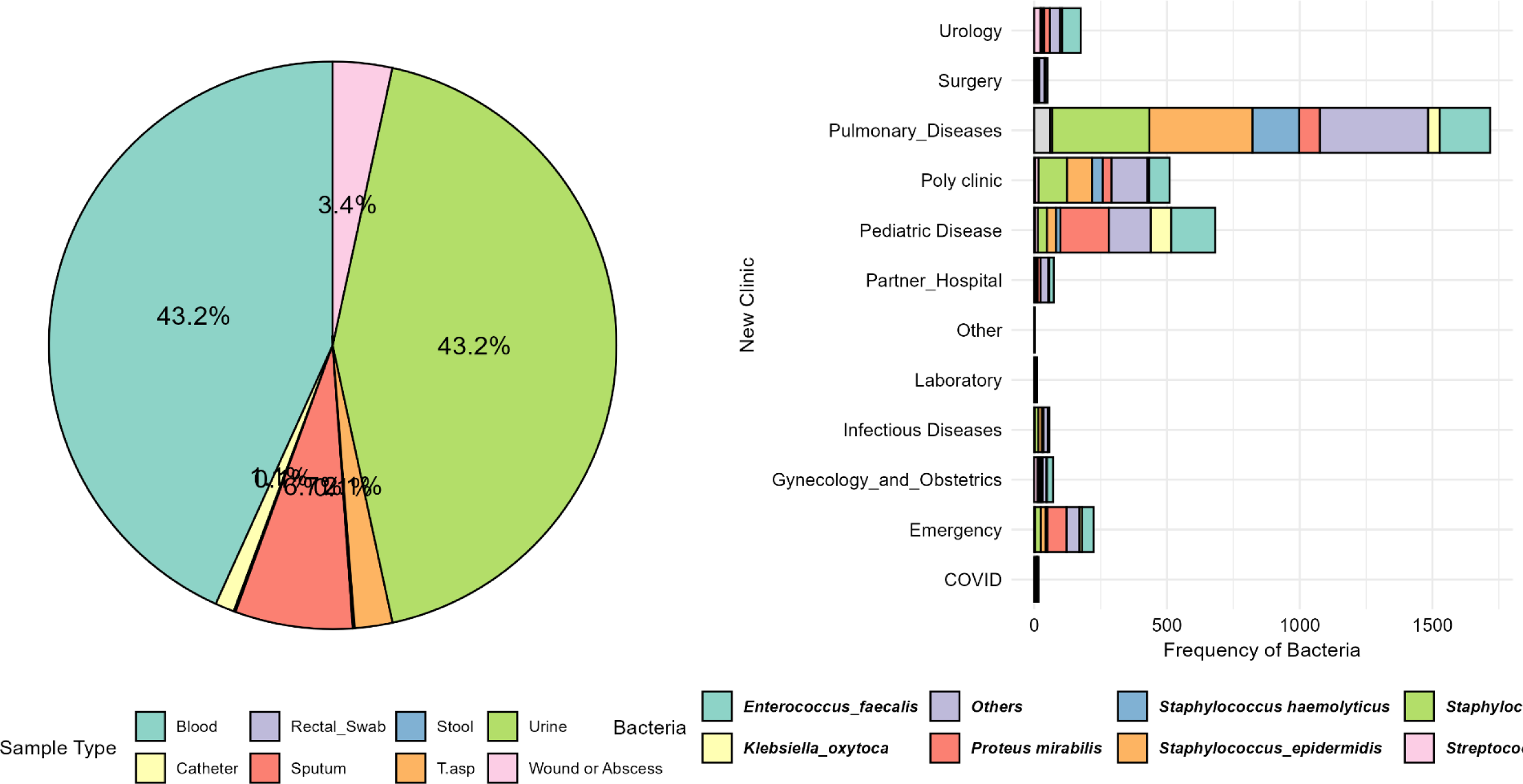

**Supplementary Table1.** Sample types and MDR category.

| Sample Type | MDR | Non_MDR | XDR |
| --- | --- | --- | --- |
| Blood | 1199 (78.11%) | 320 (20.85%) | 16 (1.04%) |
| Catheter | 40 (47.62%) | 42 (50%) | 2 (2.38%) |
| Rectal_Swab | 19 (48.72%) | 19 (48.72%) | 1 (2.56%) |
| Sputum | 63 (29.3%) | 147 (68.37%) | 5 (2.33%) |
| Stool | 17 (30.36%) | 32 (57.14%) | 7 (12.5%) |
| T.asp | 17 (29.31%) | 39 (67.24%) | 2 (3.45%) |
| Urine | 373 (25.83%) | 1050 (72.71%) | 21 (1.45%) |
| Wound or Abscess | 62 (39.24%) | 87 (55.06%) | 9 (5.7%) |

**Supplementary Table 2:**
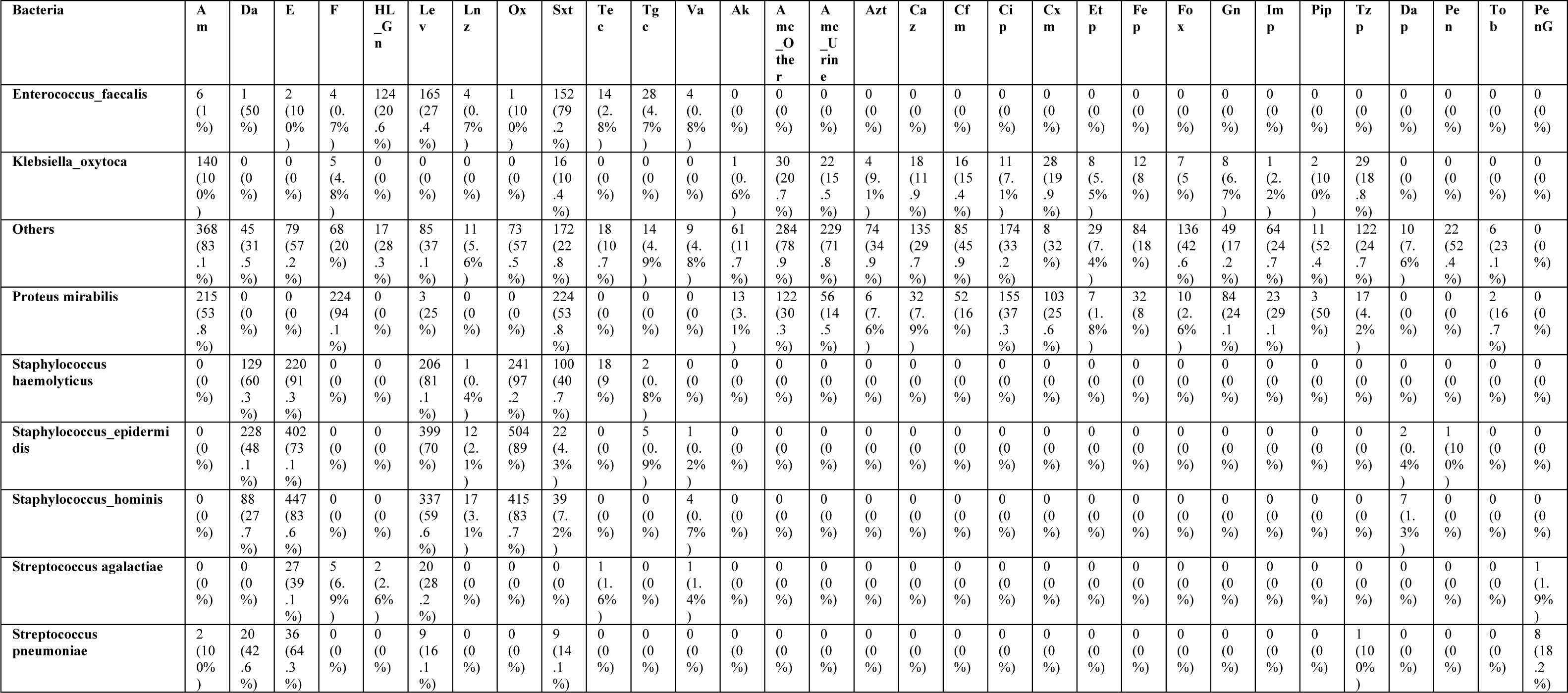
Non-ESKAPE bacteria and Antibiotics frequency and prevelance.

